# A Qualitative Descriptive Study of Motivation States for Physical Activity Among Middle-Aged Adults with Type 1 Diabetes Mellitus

**DOI:** 10.1101/2025.10.01.25336446

**Authors:** Xinyi (Vera) Wang, Garrett I. Ash, Estefania Hernandez, Matthew Stults-Kolehmainen, Stephanie A. Griggs

**Affiliations:** Case Western Reserve University, Cleveland, OH, USA; Yale University School of Medicine, West Haven, CT, USA; Yale-New Haven Hospital, New Haven, CT, USA; Emory University Nell Hodgson Woodruff School of Nursing, Atlanta, GA, USA

**Keywords:** adults, diabetes, type 1, physical activity, motivation

## Abstract

**Background:** Adults with type 1 diabetes mellitus (T1DM) have barriers to physical activity (PA), including physiological, psychological, emotional, and/or contextual. To complement prior quantitative work, we employed a qualitative study to explore momentary motivational states for movement and rest and to understand how internal and external factors shape these experiences among middle-aged adults with T1DM who had recently completed an exercise-support intervention.

**Methods:** We interviewed 23 middle-aged adults with T1DM (mean age 48, SD 11 years, 78% female, 91% non-Hispanic white, 35% overweight, and 26% obese), who recently completed an exercise intervention, using semi-structured interviews guided by the Wants and Aversions for Neuromuscular Tasks (WANT) model. This model addresses desires and aversions (i.e., fears) for movement and rest. We utilized directed content analysis to identify common themes, with both deductive and inductive coding.

**Results:** Participants exhibited simultaneous desires to move and rest, opting for activities like yoga or outdoor walks for active recovery. Competing motivations also emerged, such as the internal drive to exercise conflicting with physical fatigue or glycemic levels necessitating rest.

**Conclusions:** Our findings align with previous studies using the WANT model, underscoring the dynamic nature of motivation influenced by physiological parameters, aversive states, and recent activities. These insights gained can inform future interventions targeting psychological aspects of physical activity for this unique population.

## 1. Introduction

Diabetes management, particularly in Type 1 Diabetes Mellitus (T1DM), has increasingly recognized the pivotal role of physical activity in enhancing health outcomes. Movement and regular physical activity are associated with improved achievement of glycemic targets, cardiovascular health, and overall quality of life in individuals with T1DM (1). However, despite these benefits, many adults with T1DM struggle to engage in regular movement, necessitating a deeper understanding of the psychological, behavioral, and contextual factors that influence their motivations (2, 3).

In previous research, many environmental, social, and psychological factors have been significantly associated with physical activity behaviors in individuals with T1DM (2). For instance, access to safe exercise spaces and social support are critical determinants of physical activity behaviors (3). Psychological factors include a variety of cognitive, behavioral (e.g., readiness to change) and motivation-related variables, all of which interact in complex ways to facilitate or hinder physical activity (4).

There are many barriers that hinder physical activity among individuals with T1DM, such as employment, family responsibilities, and time constraints, which can complicate the ability to prioritize physical activity (5). Other barriers range from physiological factors, such as fear of hypoglycemia and fluctuating glucose levels, to psychological barriers, including anxiety and decreased motivation (6, 7). Fear of hypoglycemia, in particular, can severely deter individuals from engaging in activities that may cause their blood sugar levels to drop (8). Additionally, negative emotional states, such as depression and anxiety, are prevalent among individuals with T1DM and can further diminish motivation for physical activity (9, 10).

Cognitive theories of PA behavior, such as the Theory of Planned Behavior, Social Cognitive Theory, and other traditional expectancy–value models, have shown limited ability to account for variability in future PA behavior (11), and dual-process (e.g., Automatic-Reflective Theory (ART), Affective Heath Behavior Framework, Integrative Framework, model from Berry and Conroy, 2017 (12), and others) and organismic theories such as Self-Determination Theory have generally taken their place. These theories better accommodate the ideas of automaticity, urges, desires, aversions, and the corollaries of approach, withdrawal, etc. Self-Determination Theory is often highlighted because it provides a clear framework for understanding the balance between intrinsic and extrinsic motivation, as well as situational and long-term motivation, which is central to sustaining physical activity behaviors in adults with T1DM (13). Furthermore, compared with other commonly used models, such as Social Cognitive Theory or the Theory of Planned Behavior, Self-Determination Theory more directly addresses the psychological needs (autonomy, competence, relatedness) that are especially relevant to maintaining long-term engagement in physical activity while living with a demanding condition like T1DM. This framework has also been previously applied to adolescents with T1DM in prior qualitative work by Mak and colleagues, demonstrating its relevance for diabetes-specific movement experiences (14). However, motivation in the context of T1DM remains less well understood. To date, prior qualitative studies have examined attitudes, barriers, and emotional responses related to physical activity among adults with T1DM (5, 15, 16). For instance, Vlcek et al. identified “emotional responses to planning for exercise,” describing how adults with T1DM experienced shifting feelings, such as “groaning… a reluctance knowing this is going to take work,” when anticipating physical activity or managing potential glycemic consequences. Their analysis also noted that “pre- and post-exercise organization and emotional responses” shape readiness for activity and that exercise is “embedded in a broader socio-cultural context,” with preparation sometimes “negatively affect[ing] other areas of the person’s life.” (16) These findings highlight that emotions and inclinations toward activity vary across situations. However, prior work has not systematically conceptualized these fluctuations as momentary motivational states nor examined them through a theoretical model, such as the WANT framework, that is specifically designed to capture dynamic desires and aversions for movement and rest. While Self-Determination Theory provides a strong foundation for understanding psychological needs relevant to sustained physical activity, it offers limited guidance for explaining rapid, moment-to-moment motivational shifts that often occur in response to fluctuating glucose levels, physical symptoms, or safety concerns. These dynamic motivational processes are central to understanding movement and rest in T1DM and provide the rationale for incorporating additional frameworks, such as the WANT model, that account for transient motivational states.

The framework used for this study was the WANT Model (Wants and Aversions for Neuromuscular Tasks), initially developed by Stults-Kolehmainen and colleagues (17). The model includes 10 tenets (18) that describe dynamic and transitory motivational states (e.g., desires, urges, cravings, aversions) relevant to both physical activity and sedentary behaviors, the latter referring to waking low-energy activities performed in sitting or reclining postures. It provides a nuanced understanding of how individuals perceive, experience, and navigate their motivational impulses and emotions in the context of shifting physical and psychological stresses throughout the day (19). The want model has four quadrants, each named after the extreme motivational points within each quadrant.

1. High dread to move/high desire to rest: describes motivational states often associated with injury, illness, and similar conditions.
2. High dread to move/high dread to rest: describes motivational states often associated with surprise, shock, and uncertain situations,
3. High desire to move/high desire to rest: describes motivational states often associated with most daily experiences and motivational conflicts
4. High desire to move, high dread of rest: describes motivational states often associated with stressful/fight or flight experiences.

For adults with T1DM, it is useful to emphasize the ongoing shifts and interplays between wants and aversions for both movement and sedentary behaviors emanating from managing this condition. Although the WANT model was originally developed and validated in general adult samples, recent pilot work in adults with T1DM shows that transient motivation states are also highly relevant for this population. In an ecological momentary assessment study using WANT-based items along with continuous glucose monitoring (CGM) and activity data, (20) motivation to move or rest fluctuated across the day and varied with glycemic patterns, fear of hypoglycemia, aversion to hyperglycemia, and recent physical activity. These momentary states were also related to subsequent movement behavior. Together, these findings indicate that the motivational processes described by the WANT model are particularly relevant in the T1DM context, supporting its use as the guiding framework for this study.

Despite the growing body of literature on T1DM management and physical activity, significant gaps remain regarding the nuanced motivations of adults with T1DM. Much of the existing research is predominantly based on quantitative methodologies, which may overlook the variability and complexity of individual experiences. Qualitative research can provide a deeper understanding of the lived experiences of these individuals, highlighting the motivations and barriers they encounter in their daily lives (21). This study aims to address these gaps by employing a qualitative descriptive approach to explore the motivational states that influence physical activity engagement through an understanding of movement and rest among adults with T1DM. Building on the WANT framework, the purpose of this qualitative descriptive study was to elucidate internal and external factors that shape motivation states affecting physical activity among middle-aged adults with T1DM. We aimed to 1) understand perspectives and personal values related to physical activity and 2) explore experiences of motivation states related to movement and rest. Understanding influences on motivation states can provide insight into how to create supportive environments that encourage physical activity among adults living with T1DM.

## 2. Materials and Methods

### 2.1. Participants

This study is a descriptive qualitative study conducted with participants drawn from a parent clinical trial testing an exercise-support intervention for adults with T1DM. While recruitment occurred through the parent trial, the interviews and the qualitative analysis presented here were designed specifically to address the research questions guiding this study. A purposive sample of 24 adults was recruited via secure patient messaging from a regional medical group, ClinicalTrials.gov listings, and snowball sampling. Eligible participants met the following criteria: (1) Aged 30-65. This age range allowed us to capture motivational dynamics among middle-aged adults, who often balance substantial work, income, caregiving, and chronic disease responsibilities that shape “have to” and “want to” motivations. Prior work has shown that younger adults (e.g., college students) also experience motivational states, but these emerge from life pressures that are quite different from those in midlife. To avoid mixing distinct age-related patterns, we focused on middle-aged adults, whose life-stage demands align with the motivational processes examined in this study. (2) ≥1yr T1DM or other insulin deficiency diabetes (hereafter T1DM). (3) Lack of consistent engagement in structured aerobic or resistance exercise, which in the parent trial was defined as participating in fewer than one structured exercise session per week. This criterion reflected the parent study’s focus on adults with low baseline activity, (4) smartphone user, (5) English literate. and (6) had a healthcare provider who confirmed medical readiness for home-based exercise. Each participant provided the interview 1 week after completing the trial. Participants signed an IRB-approved informed consent (IRB #2000035846).

### 2.2. Study Design and Data Collection

Methods reporting follows COREQ criteria (Table S1). A qualitative descriptive approach was chosen to explore the motivations for movement and sedentarism/rest, providing an in-depth understanding while staying closely aligned with the data (22). We invited all 24 trial participants, of whom 23 (96%) accepted. Semi-structured interviews were conducted between June and October 2024 using a flexible interview guide via audio-recorded Zoom, allowing for in-depth insights into T1DM management and the motivational states for movement and rest. Interview questions, guided by the WANT model, aimed to assess desires and aversions (i.e., fears) for movement and rest (Table S2).

Participants were interviewed for an average of 28 minutes (range 14-73), and most (78%) kept their cameras on. Interviews were transcribed verbatim and reviewed for accuracy by the research team through comparison with the audio recordings, with any transcription errors corrected during this process. Participants were encouraged to reflect on past experiences where initial effort contrasted with eventual enjoyment or frustration related to healthy behaviors. Additionally, they discussed how fluctuating glucose levels influenced their motivation and fears regarding physical activity and rest. The faculty researchers included one exercise physiologist and one scientist in nursing experienced in T1DM clinical care and several advanced qualitative methods, both with postdoctoral training in T1DM behavioral science.

### 2.3. Data Analysis

Interviews were transcribed verbatim, cleaned, and checked for accuracy. A directed content analysis approach was used along with deductive (to align with the four quadrants of the WANT model) and inductive coding (23). The directed content analysis approach involved the close examination of data to identify common themes and patterns of meaning and ideas. Data analysis was conducted using ATLAS.ti (24). Interviewers were trained in qualitative interviewing. Two trained qualitative interviewers with backgrounds in anthropology, public health, and nursing conducted the interviews. They had no prior knowledge of the participants’ experience in the study nor any prior personal interaction with them.

The first step involved familiarization with the data, where transcripts were read multiple times to gain a deeper understanding of the content. A codebook was developed early in the process and revised as interviews were being conducted. The codebook ensured consistency in the coding process as it included definitions and interview questions as they relate to the theme. For example, the code ‘Barriers to Movement – Diabetes Specific’ was paired with the question “Do you find managing your diabetes gets in the way of your goals related to movement/rest?” alongside others.

Two members of the research team independently coded the transcripts using directed qualitative content analysis. Coding discrepancies were discussed and resolved through team consensus. The team refined the coding guidelines to ensure that codes were applied to comparable meaning units. After initial coding, conceptually similar codes were grouped into higher-order categories. These categories were then compared across transcripts and interpreted in relation to the WANT model and relevant prior literature on motivation and physical activity. Through this constant comparative process, the categories were refined and elevated into broader themes that captured patterned meaning across participants’ experiences. Review of memos (collected before and after interview) and reflexive journal notes were included in this process to address personal bias, assumptions, and reasons for involvement in the research topic. We used concept mapping to illustrate the themes and facilitate theme interpretation.

## 3. Results

### 3.1. Participant Characteristics

The final sample comprised 23 participants (Table 1), with a majority identifying as female (78%) and non-Hispanic White (91%). Substantial fractions were above recommended clinical thresholds for average glycemic values (57% above 7.0% or 53 mmol/mol HbA1c), blood pressure (43% above 130/85 mmHg), and/or body mass index (35% overweight, 26% obese). Regarding socioeconomic status, a majority (61%) reported a household income of $100,000 or more, while 74% earned over $60,000 per year. Most had private health insurance (87%) and automated insulin delivery (AID) systems (78%). Participants varied in their self-reported occupations and their impact on activity and rest (Table 2).

**Table 1.** Participant demographics.

| **Variables** | **Mean** | **SD** |
| --- | --- | --- |
| **Age (years)** | 48 | 11 years |
| **Duration Diabetes Diagnosis (years)** | 27 | 13 years |
| **HbA1c** |  |  |
| % | 6.8 | 0.9 |
| Mmol/mol | 51 | 10 |
| **Body mass index (kg/m<sup>2</sup>)** | 22.7<sup>a</sup><br>31.5<sup>b</sup> | 1.5<br>6.1 |
| **Systolic blood pressure (mmHg)** | 119 | 12 |
| **Diastolic blood pressure (mmHg)** | 79 | 8 |
|  | N | % |
| **Diagnosis** |  |  |
| Type 1 diabetes mellitus | 21 | 91.3 |
| Latent autoimmune diabetes of adulthood | 2 | 8.7 |
| **Sex** |  |  |
| Male | 5 | 21.7 |
| Female | 18 | 78.3 |
| **Race X Ethnicity** |  |  |
| Non-Hispanic White | 21 | 91.3 |
| Non-Hispanic Black | 1 | 4.3 |
| Hispanic White | 1 | 4.3 |
| **Marital Status** |  |  |
| Single | 3 | 13.0 |
| Married | 17 | 73.9 |
| Divorced | 2 | 8.7 |
| N/A | 1 | 4.3 |
| **Household Income<sup>c</sup>** |  |  |
| Less than \$60,000 | 1 | 4.3 |
| \$20,000 - \$39,999 | 2 | 8.7 |
| \$40,000 - \$59,999 | 2 | 8.7 |
| \$60,000 - \$79,999 | 2 | 8.7 |
| \$80,000 - \$90,999 | 1 | 4.3 |
| More than or equal to \$100,000 | 14 | 60.9 |
| **Generational Status** |  |  |
| Self and parent/s USA-born | 20 | 87.0 |
| Self USA-born, ≥1 parent outside USA | 3 | 13.0 |
| **Insurance** |  |  |
| Private | 20 | 87.0 |
| Public | 3 | 13.0 |
| **Diabetes Management** |  |  |
| Injections | 4 | 17.4 |
| Sensor-Augmented Pump | 1 | 4.3 |
| Automated Insulin Delivery (AID) | 18 | 78.3 |
| **Pump Brand** |  |  |
| Omnipod Dash | 3 | 13.0 |
| Omnipod 5 | 4 | 17.4 |
| Tandem t:slim X2 | 9 | 39.1 |
| Medtronic 770G | 3 | 13.0 |
| <sup>a</sup>N=9, <sup>b</sup>N=14<br><sup>c</sup>One participant dropped out before reaching the exit interview for this sub-study due to competing time demands. <sup>b</sup>One participant declined to respond to this question |  |  |

**Table 2.** Self-reported occupation and impact on activity and rest.

| Profession | Job Title | N | Activity Level | Disrupts Rest |
| --- | --- | --- | --- | --- |
| **Healthcare & Public Social Services** | Nurse Practitioner | 2 | Active / Sedentary | No / Yes |
|  | Ultrasound Tech | 1 | Mixed | No |
|  | Social Worker | 1 | Sedentary | Yes |
|  | Firefighter Paramedic | 1 | Active | Yes |
|  | Care Staff | 1 | Mixed | Yes |
|  | Police Officer | 1 | Mixed | Yes |
| **Education & Training** | Teacher | 2 | Active | No / Yes |
|  | School Faculty | 1 | Sedentary | No |
|  | Software Trainer | 1 | Sedentary | No |
| **Agricultural** | Arborist | 1 | Mixed | Yes |
|  | Wildlife Biologist | 1 | Mixed | No |
| **Creative Business** | Lighting Designer | 1 | Mixed | N/A |
|  | Marketing Consultant | 1 | Sedentary | Sometimes |
|  | Small Business Owner | 2 | Sedentary | No / No |
| **Miscellaneous** | Stay-At-Home Parent | 1 | Active | No |
|  | Facilities Coordinator | 1 | Mixed | No |
|  | Unemployed | 1 | Mixed | No |
|  | Retired | 1 | Mixed | No |
|  | Unknown | 1 | Mixed | N/A |

### 3.2. Key Motivation States Themes by Theme Domain

The Wants and Aversions in Neuromuscular Tasks (WANT) Model was used to describe participant movement-related behaviors and aversions (Figure 1). More specifically, the model helped to describe how participants approached or avoided (e.g., wants versus aversions) movement and/or sedentarism/rest, and the nature in which participants could vary in both states at the same moment. For example, someone could be in a state of high craving to both rest and move simultaneously when people chose to actively recover after a long day through yoga, meditation, or an outdoor walk. Another example of competing motivations is when someone has the strong internal drive to exercise every day; however, their muscles may be fatigued, or their blood sugars signal a need for rest and recovery. The following sections represent each of the four quadrants of the WANT model shown in Figure 1 and describe feelings reported by participants associated with these states, precursors to being in that state, and the behavioral outcomes of being in that state.

**FIGURE 1.**
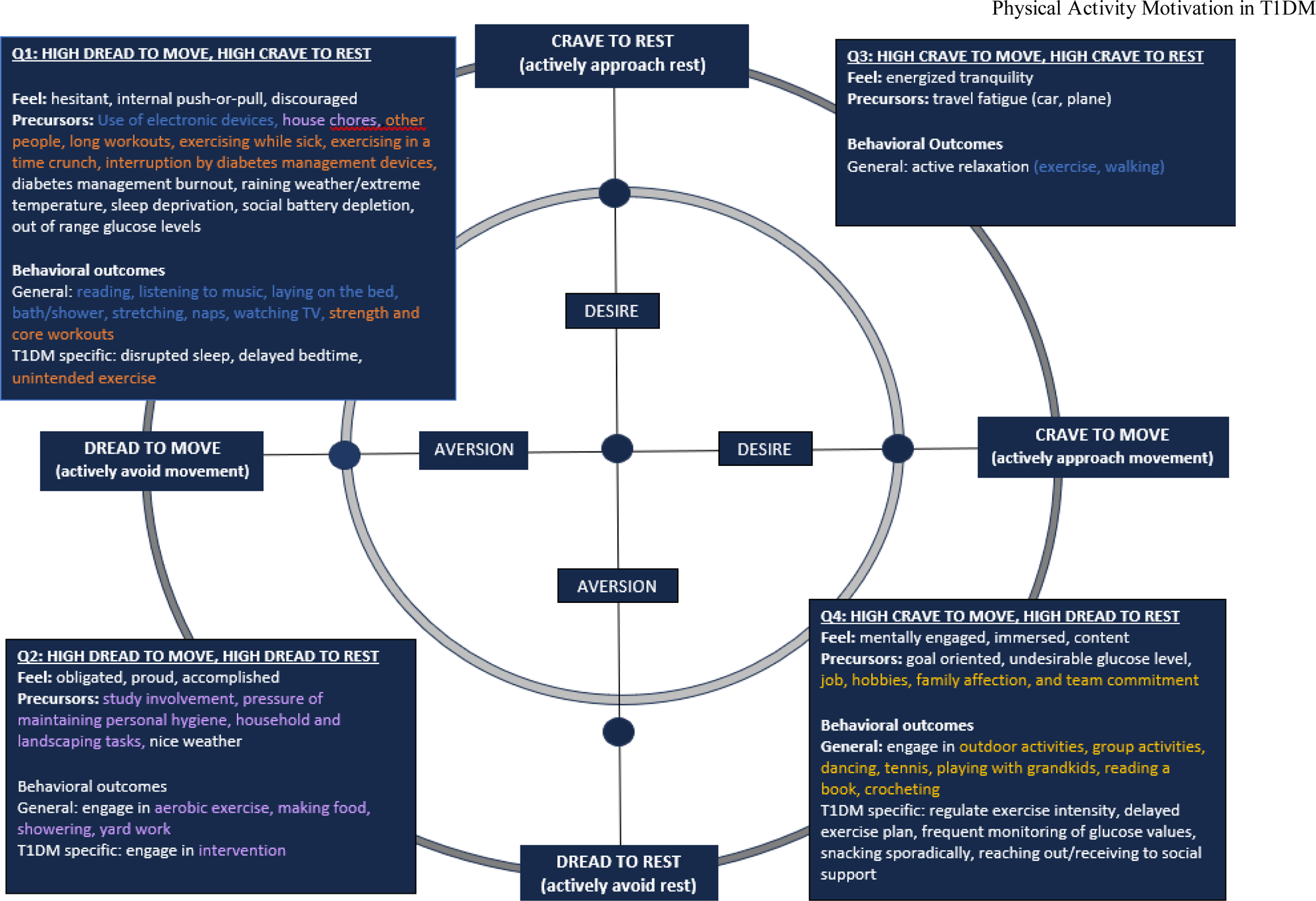
Conceptual model of analysis based on WANT Model. Highlighted colors correspond to motivational states as described by participants. Flow = Yellow; Animation = Purple; Agitation = Orange; Rest = Blue. T1DM = type 1 diabetes mellitus.

#### 3.2.1. Quadrant 1: High dread to move, high crave to rest

Participants in the current study described feelings of discouragement and hesitance to move, often experiencing a push-pull dynamic between external factors prompting movement and a strong desire to rest (i.e., motivational conflict). For instance, using electronic devices or other passive activities amplifies the urge to rest, thereby reducing motivation to initiate movement. At the same time, participants also described moments when the need for restoration led them to select light movement, such as stretching or gentle chores, as a way to feel better without committing to more demanding physical activity. Engaging in household chores served as a meaningful break, allowing participants to restore their energy. Some individuals felt uncomfortable exercising in front of others, leading to an aversion that hindered their motivation to continue movement. Additionally, workouts that exceeded their usual routines often tested their willpower, making them prone to giving up, highlighting the internal struggle in this quadrant.

Individuals expressed that when feeling unwell, their bodies typically did not function at their regular capacity, causing them to hesitate in their decision to exercise as they weigh potential benefits against drawbacks. In such instances, the body’s instinct is often to rest for a quicker recovery. However, some participants maintained that movement while feeling sick could aid in detoxification through sweating and accelerating recovery.

For participants who were parents, balancing exercise with the demands of caring for toddlers or children under time constraints, physical activity could feel burdensome. After expending energy playing with their children, their need for rest significantly increased.

Participants also mentioned that out-of-range glucose levels often hindered their ability to move. They mentally prioritized stabilizing their glucose levels to avoid further deterioration, understanding that both hypo- and hyperglycemia can disrupt normal bodily functions and necessitate rest. Conversely, some individuals indicated that they intentionally remain active as a non-medical strategy to bring their glucose levels back to a safe range, regardless of their predominant motivational state. As one participant noted, “There are times when I was not intending to do a workout, but my blood sugars are really high and stay elevated for a period, and I know that a workout will help break that cycle of high blood sugar. So, it prompts me to exercise even when I may not have wanted to or chosen to” (ID 14, Male, age 50’s, using AID for insulin delivery).

When participants were preparing for bed, malfunctioning diabetes management devices were reported to contribute to elevated glucose levels, forcing them to stay awake and troubleshoot despite feeling drowsy. For instance, one participant shared, “Sometimes you can have fluctuations in your blood sugar that make you stay up longer at night to manage it, or you might need to change your equipment because something isn’t right. That can impact your performance or sleep the next day… You’re up because your Dexcom wasn’t working, or you had a site failure, or your pump wasn’t functioning, or maybe you just ate too much…” (ID 07, Female, age 60’s, AID).

Psychologically, some participants reported burnout from managing their condition as they grapple with fluctuating glucose levels without understanding the causes. This frustration was described to drain their enthusiasm for physical activity or any movement and heightened their need for rest. Additionally, unfavorable weather, such as rainy days and extreme temperatures, significantly discouraged outdoor activities and diminished overall activity levels. Sleep deprivation led to fatigue, which in turn decreased motivation for physical activity, causing individuals to prioritize recuperation over movement.

For some, excessive time spent socializing could deplete one’s social energy, resulting in mental fatigue that diminished the desire for physical activity. As one participant expressed, “I’m an introvert, and such social situations make me more likely to want to rest” (ID 03, Female, age 40’s, sensor augmented pump [SAP]). In these moments, individuals often turn to relaxing activities like reading, listening to music, lying in bed, taking a bath or shower, stretching, napping, or watching TV as means to rest and recharge. However, in some cases of needing rest, individuals utilized strength and core workouts to relax. The simultaneous desire to rest and increased motivation for movement can lead to disrupted sleep patterns, delayed bedtimes, and unintended exercise.

#### 3.2.2. Quadrant 2: High dread to move, high dread to rest

A vast majority of participants who described this state reported often feeling a strong sense of obligation to complete tasks rather than take a break. However, once they finished, they experienced a sense of pride and accomplishment for having chosen productivity over relaxation. Some individuals, who did not have a prior exercise routine, felt compelled to stick to the study’s exercise plan, and, although they may have resisted at first, they found gratification and enjoyment in the process. This was particularly noted during intervention exercises, where participants expressed that their initial reluctance transformed into enthusiasm. As one participant shared, “One day I didn’t really want to go in the pool because it looked really dark and cloudy and wasn’t that warm, but I was on the phone with [STUDY MEMBER], and I kind of forced myself. And then once I was in, I was able to stay in for a while” (ID 12, Female, age 50’s, AID).

Similarly, some participants viewed taking a shower as a chore, motivated by the pressure to maintain personal hygiene. Yet, once they started, they often emerged feeling refreshed and rejuvenated. The same theme persisted with yard work; despite initial resistance to maintaining their household and landscape, individuals pushed themselves to engage in these activities. After completing the work, they were rewarded with the sight of a well-kept yard. Positive weather conditions played a significant role, often prompting participants to prioritize outdoor chores over resting indoors. One participant remarked, “I’m going to have to do yard work, and I just don’t want to because I’m tired but it’s a beautiful day out so it urges me to not want to rest because it’s so nice outside, and I want to go do something” (ID 11, Male, age 30’s, AID).

In cases where individuals struggled to motivate themselves, they often began with aerobic activities. Gradually, as they engaged more fully, they began to take pride in their progress and the improvement in their physical condition. Furthermore, while balanced meals are crucial for managing T1DM, participants did not always cook out of desire but rather necessity. They opted to spend time preparing healthy meals to regulate their glucose levels, rather than skipping meals or choosing easier alternatives. Ironically, many found enjoyment in the cooking process once they started, revealing how initial aversions can shift to positive experiences.

#### 3.2.3. Quadrant 3: High crave to move, high crave to rest

Individuals often described themselves in a state of energized tranquility when their bodies signaled a need for recovery, prompting a desire to balance this need with dynamic activities. The craving for rest could lead to an instinctive counteraction, where individuals engaged in movement, thereby fostering a harmonious flow between action and relaxation. For instance, during long car or air travels, discomfort could arise from narrow spaces, prolonged sitting, and insufficient opportunities for quality rest, culminating in fatigue. The confined environment placed a strain on muscles and increased feelings of restlessness. These factors combined to create stiffness and fatigue, amplifying the urge to move to alleviate tension and restore comfort.

As one participant articulated, “When we were driving for three days straight, after about seven hours in the car, you’re like, ‘I need to stop driving,’ but it’s an hour and a half until we reach our destination, and I’m like, ‘I need to move.’ We are stopping right now because I have to move. I have to get out and walk. I have to do something. I gotta jog. I gotta do some jumping jacks. I need to do something because I’m going to go crazy in the last 90 minutes” (ID 06, Female, age 50’s, AID).

Similarly, after engaging in strenuous exercise, individuals often resorted to active relaxation techniques, such as stretching or walking, to meet their body’s recovery needs. One participant noted, “After exercise, I usually rest and recover by walking” (ID 02, Female, age 40’s, AID). This interplay between the desire to move and the need for rest illustrates the complex dynamic encapsulated by the Wants and Aversions in Neuromuscular Tasks (WANT) Model, highlighting how individuals navigate competing desires simultaneously.

#### 3.2.4. Quadrant 4: High crave to move, high dread to rest

Several participants articulated the experienced heightened sense of mental engagement when participating in enjoyable physical activities, leading to a reluctance to rest. The satisfaction derived from completing these activities fostered a sense of contentment. For those who described themselves as naturally competitive, the effort needed to achieve movement goals was a significant source of pleasure. When setting such goals, motivation stemmed from the desire to reach new heights. People who took pleasure in their work frequently expressed enjoyment during their tasks, making them less inclined to seek rest. Engaging in personal hobbies was also reported to induce a flow state, wherein individuals become fully absorbed in the activity without perceiving a need for rest, as the enjoyment and focus they derive sustained their energy levels.

Quality time spent being active with family was described as creating strong emotional connections, reinforcing family bonds, and cultivating a sense of contentment. This positive interaction spurred individuals to continue moving, buoyed by the energy of shared experiences rather than seeking out moments of rest. Team leaders in sports reported feeling a commitment to their peers, motivating them to attend practices and participate actively rather than opting for rest. Furthermore, a sense of responsibility to keep teammates engaged and serve as a role model was described to enhance their motivation to stay active.

In the context of managing blood sugar levels, participants oftentimes gravitated toward moderate-intensity exercise as a practical means to lower glucose levels, opting for movement over rest to address their needs. As one participant expressed, “If I could somehow have the freedom, which I can’t, my diabetes doesn’t let me, to exhaust myself with physical activity on a daily basis, I would” (ID 09, Female, age 60’s, HCL). Another noted, “I’m conscious of [my sugars] all the time. If they’re on the lower side, I probably wouldn’t start with vigorous exercise or workout or anything” (ID 08, Female, age 30’s, using injections for insulin delivery).

To achieve feelings of contentment and enjoyment, individuals typically chose activities that promote movement and interaction rather than rest. Such activities encompassed outdoor excursions, group engagement, dancing, playing tennis, interacting with grandchildren, or even pursuing hobbies like reading or crocheting. However, before embarking on physical activities, participants’ glucose levels could influence the intensity of their exercise, which could at times cause delays or hinder their movement goals. They often required periods of rest to recheck and adjust their glucose levels to ensure they are within a safe range before resuming physical activity.

“Every once in a while, I’d run into that fact of where I’d have a spike in my blood sugar to be high, and when I want to do a workout, which I know is not the best. And then it’s like, I got to wait for it to catch up” (ID 11, Male, age 30’s, AID). Similarly, another participant shared, “Sometimes I might feel an urge or need to do a physical activity later than I would have wanted, because my blood sugars are high” (ID 14, Male, age 30’s, AID).

Given the significant impact of movement on glucose levels, participants reported the need to consistently monitor their levels throughout activities, which often disrupted their overall experience. These necessary adjustments lessened enjoyment and reduced feelings of contentment as activities came to an end. “When I was doing the evening walks, there were several days where I had dinner a little bit later than I planned, and I still had so much insulin on board that I didn’t think it was a good idea to go out and walk away from home because of the insulin on board” (ID 10, Female, age 40’s, AID). Another participant recounted, “We were swimming and I couldn’t [continue] because I was starting to go low” (ID 18, Female, age 50’s, AID).

## 4. Discussion

The current study aimed to explore motivation states for physical activity and sedentary behaviors among middle-aged adults living with Type 1 Diabetes (T1DM) using the WANT Model framework (18). Understanding the dynamic interplay of psychological factors influencing movement and rest among adults with T1DM is crucial for developing effective interventions. This discussion highlights the implications of our findings, the strengths and limitations of our study design, and recommendations for future research.

Our findings illustrate how the interaction between desires (e.g., urges, cravings) and aversions (e.g., dread, fear) informs participants’ choices about engaging in movement versus resting behaviors. Additionally, our results align with a mixed methods study of 17 undergraduate students revealing three similarities: (1) there was a dynamic nature of motivation noted where a fluctuating nature of motivation states was found, frequently with rapid changes in desires to move and rest, (2) the influence of external factors (e.g., pressure of maintaining personal hygiene) was significant, and (3) strong urges manifested from the deprivation of both movement and rest (17). These patterns are consistent with the idea that challenging or uncomfortable conditions can make people more likely to choose lower-energy activities. However, the nuances introduced in the T1DM context highlight the unique challenges and conflicts individuals face when managing this complex condition, such as having a low blood sugar level, preventing exercise from occurring simultaneously with an urge to exercise. Further, the current study focused on middle-aged adults who were grappling with a complex chronic condition, and these demographic differences may have led to varied motivational experiences due to differences in life stage, responsibilities, and health management. This suggests that future interventions or research would benefit from considering these complex influences on motivation alongside physiological states to better address the needs of diverse populations.

The most hypothesized diseases to be impacted would be movement-related disorders, and a scoping search found language consistent with the WANT model in 95 studies of such conditions (25) (Table S3). But available evidence has also suggested more complex pathways involved in chronic psychiatric disease, such as in anorexia nervosa, where the drive to be active is propelled by body-conscious ideations (26), or those involved with state anxiety and other mental disorders (27, 28). Meanwhile, studies of healthy populations have generated evidence that momentary placement within the WANT model is associated with stimuli similar to what we observed in the present study including energy availability (a correlate of glucose levels in T1DM), aversive valence states (akin to anticipation of unpleasant glucose trends), time of day, and recent physical activity or sedentary time (29). Elevated glucose may therefore influence motivation by signaling increased energy availability, or, as participants suggested, by prompting movement to maintain or restore glucose to target ranges with less insulin.

The findings of this study contribute to the growing body of literature emphasizing the profound connection between physiological states, such as glucose levels, psychological outcomes, particularly motivation, and behavior. Existing research has demonstrated that individuals with T1DM often experience fluctuations in mood and motivation due to shifts in their blood glucose levels. For instance, studies have illustrated how both hyperglycemia and hypoglycemia can lead not only to physical symptoms but also to cognitive impairments and aversions that affect decision-making processes (30, 31).

Furthermore, aligning with findings from a study by McCarthy et al. (32), which pointed out the behavioral impacts of diabetes management on physical activity, the current study reinforces the argument that glucose fluctuations not only influence physical health but also the psychosocial dimensions of living with diabetes. This suggests a critical gap in current interventions, which often focus primarily on the physiological aspects of diabetes management, neglecting the psychological and motivational layers that are equally crucial for effective self-management. A number of studies have addressed the fear of hypoglycemia by educational or cognitive behavioral approaches (33, 34), but studies like the current one reveal the complexity of physical activity behavior and determinants beyond fear of hypoglycemia (35, 36).

Overall, the emphasis on tailored interventions that address these motivational nuances aligns with recommendations from researchers advocating for holistic, patient-centered approaches in diabetes care (37). Such interventions could enhance adherence to recommended physical activity guidelines, ultimately improving both psychological well-being and physical health outcomes for individuals with T1DM.

When interpreting the results of this study, several limitations must be acknowledged. We enrolled individuals at the end of an intervention designed to address inconsistent exercise, leaving the possibility that findings do not apply to individuals with long-term high or low activity levels. Our sample, like the adult cohort of the type 1 diabetes exchange (38), had limited racial or ethnic diversity, though it was diverse regarding HbA1c, blood pressure, anthropometrics, and occupational physical activity (Table 1). Additionally, the qualitative nature of the study may introduce biases, as participants could have altered their responses due to social desirability. To address these limitations, future research should focus on larger, longitudinal qualitative and quantitative studies to examine the stability of motivation states over time and their long-term impacts on movement and sedentary behaviors.

Despite these limitations, it is important to recognize the strengths of this study. The qualitative methodology employed allowed for an in-depth exploration of participants’ lived experiences, with semi-structured interviews yielding rich, nuanced data that captures the complexity of motivation states. Furthermore, to our knowledge, this is the first study to explore motivation states in middle-aged adults with T1DM using the WANT model. This approach not only sheds light on the motivational dynamics within this population but also lays the groundwork for further research and targeted interventions tailored to their specific needs and desires.

Another theme that emerged from the data, though not explicitly delineated in the paper, was the recurring language of “have to” (obligation) reflecting a sense of necessity in participants’ motivation. This dimension permeated throughout the results and may represent an important complement to the “want to/don’t want to” motivational perceptions emphasized in the WANT model. For future studies, it may be useful to explore how a sense of obligation interacts with “want to” or “don’t want to” motivations to influence behavior and how it’s further modulated by motivational conflicts (e.g., “want to move” vs concurrent “want to rest”), a finding corroborated in our previous qualitative study with 17 adults without any chronic disease (17).

Moving forward, future researchers should focus on larger, longitudinal studies to examine the stability of motivation states over time and their long-term impacts on movement and sedentary behaviors/rest. Additionally, investigating how varying motivational states can inform targeted intervention designs aimed at improving physical activity among individuals with T1DM is essential. It is also crucial to explore the role of healthcare providers in facilitating conversations about motivation and offering ongoing support tailored to individual experiences and challenges. By addressing both the limitations and strengths of this study, we can pave the way for more effective strategies to enhance T1DM self-management support.

## Declarations

The authors have no conflicts of interest to disclose.

Garrett Ash and Stephanie Griggs report financial support was provided by the National Institutes of Health. Garrett Ash reports a relationship with American Heart Association Inc that includes: funding grants. Garrett Ash reports a relationship with The Patterson Foundation that includes: funding grants. Garrett Ash reports a relationship with Calm.com that includes: professional services for a nominal fee. Stephanie Griggs reports a relationship with the National Institutes of Health that includes: funding grants. Other authors declare that they have no known competing financial interests or personal relationships that could have appeared to influence the work reported in this paper.

## Ethics approval and consent to participate

Participants signed a Yale IRB-approved informed consent (#2000035846).

## Consent for publication

Yes.

## Availability of data and materials

The data will be available immediately following publication, with no end dates. The data will be shared with researchers who provide a methodologically sound proposal to achieve the aims of the approved proposal. Data will be truncated to meet the proposal aims without compromising participant confidentiality. The proposals should be directed to Dr. Garrett Ash at Yale University. To gain access, data requesters must sign a data access agreement.

## Authors’ contributions (CRediT Statement)

GIA, conceptualization, data curation, funding acquisition, investigation, project administration, resources, writing – reviewing and editing. EH, data curation, formal analysis, validation, visualization, writing – original draft. XW, formal analysis, validation, visualization, writing – original draft. MSK, validation, writing – review and editing. SG, conceptualization, data curation, formal analysis, investigation, methodology, project administration, supervision, validation, visualization, writing – original draft.

## Acknowledgments

The authors acknowledge that this abstract was presented at the American Diabetes Association (ADA) Scientific Sessions, [2025].

## Glossary

HbA1c: hemoglobin A1c
T1DM: type 1 diabetes mellitus
WANT: Wants and Aversions for Neuromuscular Tasks model.

## Supplemental Material

**Table S1.** COREQ Checklist.

**Table S2.** Interview Guide.

**Table S3.** Literature Review.

